# Theory of radiologist interaction with instant messaging decision support tools: a sequential-explanatory study

**DOI:** 10.1101/2023.06.15.23291470

**Authors:** John Lee Burns, Judy Wawira Gichoya, Marc D Kohli, Josette Jones, Saptarshi Purkayastha

**Author notes:** Corresponding author (A1).

## Abstract

Radiology specific clinical decision support systems (CDSS) and artificial intelligence are poorly integrated into the radiologist workflow. Current research and development efforts of radiology CDSS focus on 5 main interventions, based around exam centric time points– at time of radiology exam ordering, after image acquisition, intra-report support, post-report analysis, and radiology workflow adjacent. We review the literature surrounding CDSS tools in these time points, requirements for CDSS workflow augmentation, and technologies that support clinician to computer workflow augmentation.

We develop a theory of radiologist-decision tool interaction using a sequential explanatory study design. The study consists of 2 phases, the first a quantitative survey and the second a qualitative interview study. The phase 1 survey identifies differences between average users and radiologist users in software interventions using the User Acceptance of Information Technology: Toward a Unified View (UTAUT) framework. Phase 2 semi-structured interviews provide narratives on why these differences are found. To build this theory, we propose a novel solution called Radibot - a conversational agent capable of engaging clinicians with CDSS as an assistant using existing instant messaging systems supporting hospital communications. This work contributes an understanding of how radiologist-users differ from the average user and can be utilized by software developers to increase satisfaction of CDSS tools within radiology.

**Author Summary:** There is a need for human-machine interfaces between radiologists and clinical decision support systems (CDSS). Within the variety of systems radiologists interact with, there is no best fit for CDSS presented in the literature. After reviewing current literature surrounding CDSS use in healthcare and radiology, we propose a novel solution - a conversational agent capable of engaging clinicians as a team member using existing instant messaging systems supporting hospital communications.

We test the acceptance of this intervention using the User Acceptance of Information Technology: Toward a Unified View (UTAUT) framework in survey and interview formats. Within our sample group, we found that radiologists have a high intent to use and a positive attitude towards this intervention. Our sample of radiologists deviated from the standard user UTAUT expects, suggesting that radiologist’s acceptance of software tools differs from the standard user. This work builds a theory of radiologist-decision support tool interaction that may be useful for software developers and systems integrators.

## 1 Introduction

Radiology domain-specific clinical decision support systems (CDSS) applications are poorly integrated into the radiologist workflow (1). In 2017, Dreyer and Geis described a transition in radiology moving towards integrating Artificial Intelligence (AI) into the radiologist workflow. "In the past, radiology was reinvented as a fully digital domain when new tools, PACS and digital modalities, were combined with new workflows and environments that took advantage of the tools. Similarly, a new cognitive radiology domain will appear when AI tools combine with new human-plus-computer workflows and environments." They describe the concept of a "Centaur Radiologist" as a physician utilizing AI-augmented CDSS workflows to increase efficiency (2). We expand this term as “future radiologist,” inclusive of non-AI techniques in CDSS.

However, the future radiologist will not happen if the tools are poorly integrated, with cumbersome human-computer interfaces (3). Deliberate and sustained effort by using inter-disciplinary knowledge from human-centered computing, psychology, cognitive sciences, and medicine is required to build CDSS for the future radiologist (4). In this work we create a basis of knowledge in the theory of radiologist-decision tool interaction using a sequential explanatory study design. The study consists of 2 phases, the first a quantitative survey and the second a qualitative interview study. The phase 1 survey identifies differences between average users and radiologist users in software interventions using the User Acceptance of Information Technology: Toward a Unified View (UTAUT) framework (5). Phase 2 semi-structured interviews provide narratives on why these differences are found. To build this theory, we propose a novel solution called *Radibot* - a conversational agent (CA) capable of engaging clinicians with CDSS as an assistant using existing instant messaging (IM) systems supporting hospital communications. This work contributes an understanding of how radiologist-users differ from the average user and can be utilized by software developers to increase satisfaction of CDSS tools within radiology.

### 1.1 Background

We expect that the future radiologist will routinely interact with CDSS at each stage of their workflow. We designed Radibot for diagnostic radiologists, with interventions at each of the following time-points: after image acquisition, during report creation, after report creation, and between studies. A brief overview of existing interventions in each time point follows.

- After Image Acquisition - radiologists combine a variety of data to make interpretations of images. Interventions include Computer-Aided Detection (CAD), Computer-Aided Diagnosis (CADx), and patient history/metadata presentation. These interventions generally function within the Picture Archiving and Communication System (PACS), though some will interface with the Radiology Information System (RIS), Voice Recognition system (VR), or in an external client (6-16).
- During Report Creation – these interventions surround embedding evidence-based guideline processes during dictation and are found within VR. Guidelines are navigated using drill-through commands or natural language processing (NLP) of the dictation to generate report text (17, 18).
- After Report Creation - In most RIS, reports are stored as unstructured text. Interventions in post-report analysis include extracting categorical data, automating radiologist-clinician communication, and quality improvement systems. By generating summative report metadata, these interventions enable context-switching and reduce fatigue when a radiologist is asked to return to a finished report (19-31).
- Between Studies – existing adjacent to radiologist workflow, these interventions influence decision making at an individual or business level and consist of workflow-prioritization, management, and feedback tools. These tools utilize metadata found in Health Level 7 (HL7) or Digital Imaging and Communications in Medicine (DICOM) messages. Users interface with them outside of clinical systems, IE. web dashboards, or they are integrated into PACS/RIS/VR presentation layers (32-39).

Diagnostic radiologist’s clinical work is fully completed using systems, including PACS, RIS, and VR, with every interaction being digitally augmented (40). Given the fully digital clinical workflows, radiology specific CDSS implementations are uniquely positioned to provide support and affect change. Radiology specific guidelines for "advisor systems" were laid out by Teather et al. in 1985, while Khorasani in 2006 provides features for the development of clinical decision support systems (41, 42). Outside of radiology, CDSS are built following the Ten Commandments for Effective Clinical Decision Support: Making the Practice of Evidence-Based Medicine a Reality (43). Commandments 2, 3, 7, 10, and 1 – anticipate needs, fit into user workflow, simple interventions, knowledge system maintenance, and speed – appear with a higher frequency when aligned with radiology specific guidance. An alignment of the general CDSS and radiology specific CDSS guidelines are found in table 1. Differences in CDSS priorities underscore the need for more research in this area and are mapped to UTAUT concepts and the hypotheses for phase 1.

**Table 1.** Commandments compared to published considerations.

| Commandments<sup>a</sup>/Considerations | Speed is Everything | Anticipate Needs and Deliver in Real Time | Fit Into the Users Workflow | Little Things Can Make a Big Difference | Recognize that Physicians Will Strongly Resist Stopping | Changing Direction is Easier than Stopping | Simple Interventions Work Best | Ask for Additional Information Only When You Really Need It | Monitor Impact, Get Feedback, and Respond | Manage and Maintain Your Knowledge-Based Systems |
| --- | --- | --- | --- | --- | --- | --- | --- | --- | --- | --- |
| "The system must follow the usual working practices of the clinician and must not appear to usurp his/her position." <sup>b</sup> |  |  | X |  | X |  |  |  |  |  |
| "The system must have a good, adaptable user interface which uses clinical terminology, and is able to give help on demand." <sup>b</sup> | X | X | X |  |  |  | X |  |  |  |
| "Diagnostic advice (if sought) must be 'calibrated' so that uncertainties output by the system may be interpreted in terms of the incidence of errors. Explanation/justification of conclusions should be available on demand." <sup>b</sup> |  | X |  |  |  |  |  | X |  | X |
| "For the system to be used, rather than just usable, it must offer more than just simple diagnostic advice, and these other facilities should be available independently of diagnostic advice." <sup>b</sup> |  | X | X |  |  |  |  |  |  | X |
| Time sensitivity, with a preference for real time CDSS. <sup>c</sup> | X | X |  |  |  |  |  |  |  |  |
| Brevity in providing salient information to answer the clinical question asked and link to evidence. <sup>c</sup> |  |  | X | X |  |  | X |  |  |  |
| Recommend action and provide descriptive reasoning and actionable interventions <sup>c</sup> |  |  |  |  |  | X | X |  |  |  |
| CDSS must use up to date evidence-based medicine <sup>c</sup> |  |  |  |  |  |  |  |  |  | X |
| CDSS should improve productivity, efficiency, quality, and safety. <sup>c</sup> |  |  |  |  |  |  |  |  | X |  |

<sup>a</sup> 10 commandments from Bates et al. (43).
<sup>b</sup> Considerations quoted from Teather et al. (41)
<sup>c</sup> Considerations paraphrased from Khorasani (42),

### 1.2 Instant Messaging and Conversational Agents in Healthcare

IM is found throughout the healthcare enterprise, including in disease management, patient-clinician interactions, medical education, among patient populations and workforce members for extra-clinical activities. IM can be inclusive of voice, video calling, and file sharing (44). Extra-clinically, IM tools facilitate socialization, catharsis, and professional connectiveness functionalities when applied in clinical settings (45, 46). IM is asynchronous and short-form, leading to advantages over other communication methods, particularly in the area of articulation work - answering medical questions, coordinating logistics, addressing social information for patients, and querying staff/equipment locations or status (47). IM is integrated into many PACS, RIS, and VR, serving many purposes within radiology including care discussions and facilitating remote tele-radiology communications (29, 48-58).

CA, or chatbots, are natural language human-machine interfaces. CA can apply 4 methods for negotiating user interactions: immediate, negotiated, mediated, and scheduled (59). Consumer health care CA are currently scheduling appointments, providing basic symptom identification and recommendation, and assisting with long term care such as sensor monitoring/alerting and medication reminders (60). Most healthcare CA are built for patients (interview, data collection, or telemonitoring), while clinician focused CA are designed around data collection (61). Other efforts in clinician focused CA include interpreting spoken language into clinical facts and drug interaction/alternative drug recommendation systems (62-64). IM impact on task completion is not fully understood, especially in the context of automated IM interventions. There is evidence that non-relevant messages can increase or reduce task completion times depending on the message initiator; at a cost of quality of the task output (65). Disruptiveness of IM specific interventions is reduced when IM are relevant to the task being completed or if delivered at time-points that fit the users workflow (66). IM interactions among a professional workforce are found to support task completion, accuracy, and quality of outcomes (65).

## 2 Methods

### 2.1 Population

Our study population consists of 2 sets of radiologists – attendings and trainees at a large academic health system. The attendings set is a subset of the approximately 112 radiologist faculty at a teaching hospital system. The trainee’s set is a subset of the 62 residents/fellows within the same system. Our population is acquired through convenience sampling. Of 174 possible participants, 98 responded affirmative that they would complete the survey and 3 that they did not want to participate. 39 participants responded that they would complete an interview and 11 responded that they would participate in the survey but not the interview. In total, 94 surveys were submitted, and 23 interviews were conducted.

### 2.2 Survey

An electronic survey was created using Qualtrics (67), we collect population composition and quantitative data surrounding intervention feasibility, usability, and acceptance. Within the UTAUT framework we focused on behavioral intention to use the system (BI), attitude toward using the technology (AF), effort expectancy (EOU), performance expectancy (PE) and anxiety (ANX). We chose to not utilize questions in social influence, facilitating conditions, and self-efficacy due to applicability to a prospective study of a tool not yet implemented in practice. A full listing of UTAUT questions by construct and factor are found on EDUTECH’s Wiki (68). Due to respondent time constraints we chose to utilize 12 of 19 questions in the chosen constructs, with each construct having at least 2 questions asked. Questions were eliminated if they were not relevant to a system that does not yet exist (Example: Working with the system is fun).

Other frameworks exist for testing usability and user experience for software design. However, UTAUT is unique in the number of constructs it can capture quickly. Measures like the System Usability Scale or Technology Acceptance Model can capture intent to use, but do not create the linkages to moderating factors of interest. Contrasting the UTAUT concepts with the CDSS commandments, we create the following links:

- Performance Expectancy
  - 1. Speed is everything
  - 2. Anticipate Needs and Deliver in Real Time
  - 5. Recognize that physicians will strongly resist stopping
  - 7. Simple interventions work best
- Effort Expectancy
  - 3. Fit into the user’s workflow
  - 4. Little things can make a big difference
  - 6. Changing Direction is Easier than Stopping
  - 8. Ask for Additional Information Only When You Really Need It
- Anxiety
  - 5. Recognize that physicians will strongly resist stopping

The survey in full is included in the S1 Appendix A.1. Fig 1 highlights the intervention and proposed capabilities.

**Fig 1.**
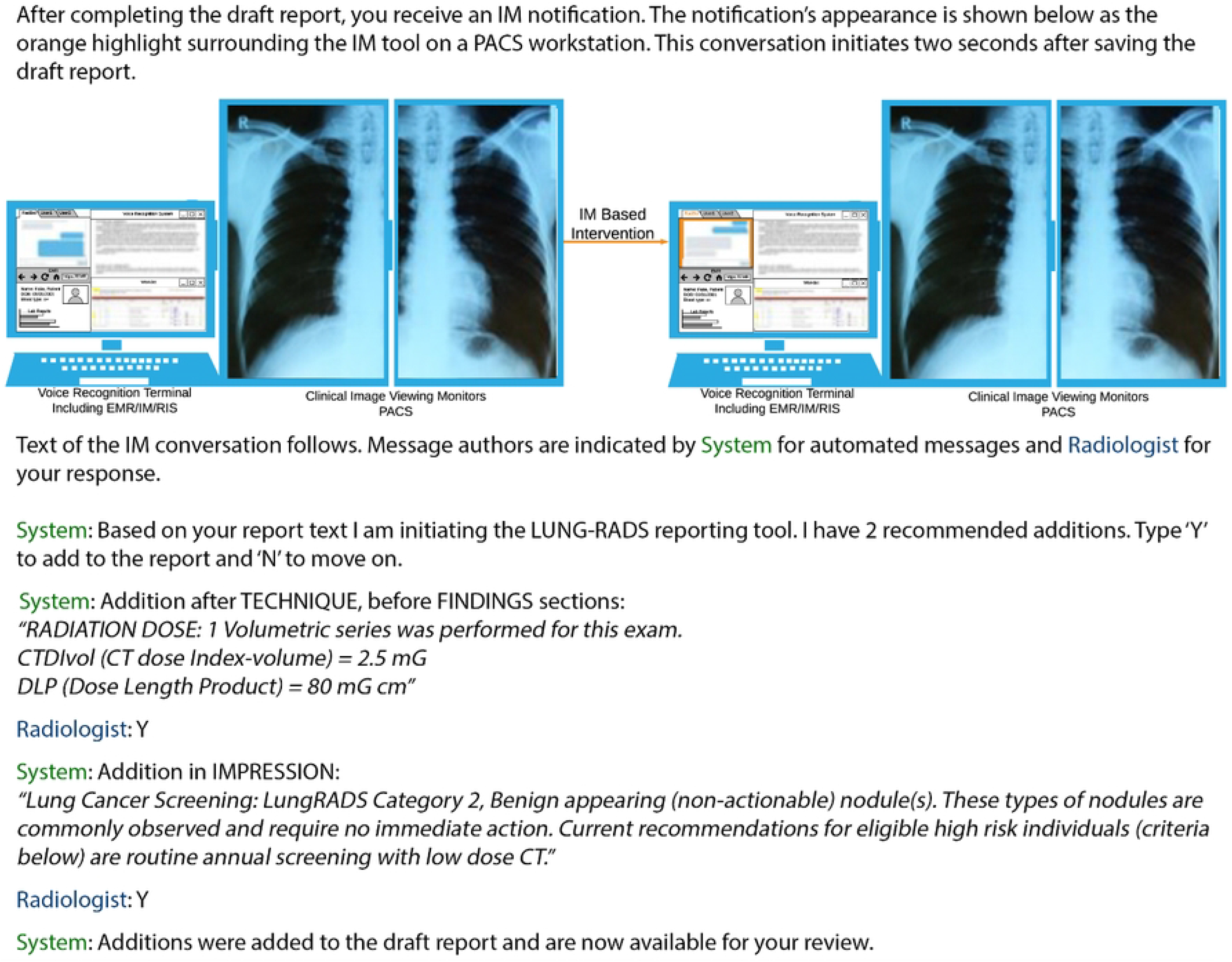
Sample PACS workstation before/after IM based intervention, and details of intervention presented to survey takers. Source images for Lung X-Ray (69), Report (70), and IM transaction (71). LUNG-RAD scenario and output text (72).

### 2.3 Interview

Using the research statements developed with the survey (S1 Appendix A.5), we generated hypothesis and began developing the semi-structured interviews. As we did not have a working system, we prototyped 5 interventions and created video examples of each to use during the interview. Fig 2 highlights what these videos looked like during a demo. The videos highlighted interventions during each workflow time point in the following ways:

- After Image Acquisition
  - Video 1 – Radibot identifies potential for 3d reconstruction, asks radiologist permission to process, and then suggests the correct VR template.
  - During Report Creation
  - Video 2 – Radiologist engages Radibot to query the Electronic Medical Record (EMR) for cardiac risk factors. Radibot performs this query as the radiologist returns to reviewing images, then returns all risk factors that meet these criteria.
  - Video 3 – Radibot identifies VR dictation of left adrenal nodule then engages radiologist in stepping through adrenal nodule flow chart. Completion of the flowchart inserts guideline recommend text and citation into report.
  - After Report Creation
  - Video 4 – Based on text of report, Radibot engages radiologist for follow up communication.
- Between Studies
  - Video 5 – Radibot presents possible studies for radiologists to engage with, removing the need to navigate the worklist. Includes suggestions of cross coverage of busier worklists and high priority studies.

**Fig 2.**
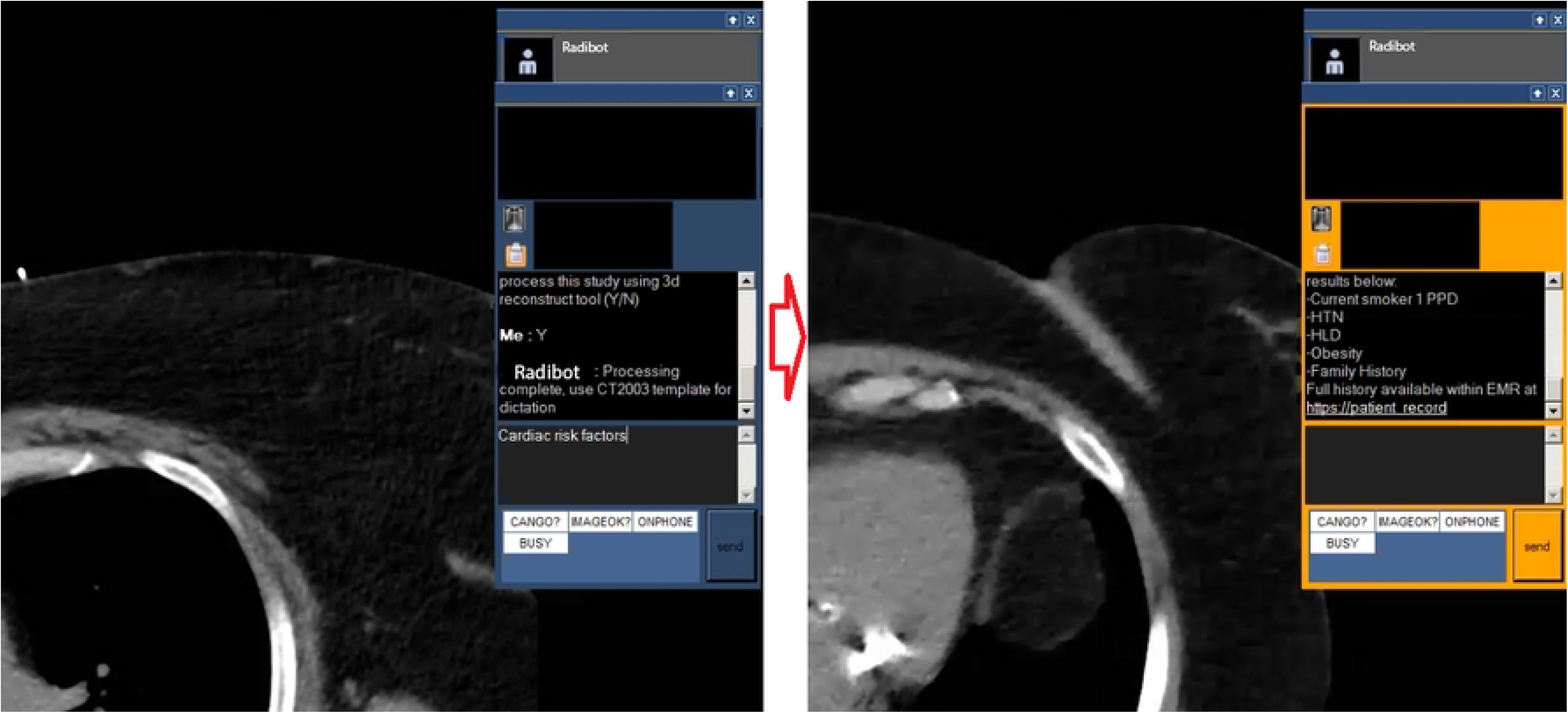
Capture from Video 2 highlighting radiologist query and Radibot response.

An interview guide was created (S1 Appendix B.1) following the UTAUT framework. The guide begins with video 1, loops through each video asking the same questions, then has a set of questions after all videos have completed. A portable interview setup was created consisting of one laptop, a 4k portrait monitor mimicking a diagnostic monitor, and a microphone for collecting audio. Interviews occurred in offices/conference rooms located near interview candidates normal work locations. Subjects were presented with consent and informed that no names would be utilized during the interview for confidentiality. Zoom was utilized to record the screen and interview narrative to the laptop (73).

39 survey participants responded that they would complete an interview. 23 interviews occurred before the research team agreed that response saturation was achieved. Interviews were transcribed using Otter.AI, then a research assistant and study team member reviewed each video separately and corrected any transcription errors (74). Transcriptions were downloaded in docx format, then loaded into ATLAS.ti 9.0.19.0 for qualitative analysis. The study team created labels for text analysis (S1 Appendix B.2) and linked these by semantic domain (UTAUT construct). 2 research assistants were hired and trained by the study team to annotate interview text using ATLAS.ti. The research assistants separately annotated interview 1, then the study team reviewed and provided additional guidance. They then separately annotated the remaining interview narratives, and the annotated narratives were merged, and inter-rater agreement is measured. Because semantic domains are established and we did not segment quotes in advance, Krippendorff’s CU Alpha is utilized to measure semantic domain agreement by quote. An overall agreement level of α ≥ .8 is set for all documents (75).

## 3 Results

### 3.1 Survey Data Analysis

Resulting data was downloaded from Qualtrics in Comma Separated Values (CSV) format and analyzed using Excel. Irrelevant metadata fields were removed. A total of 88 survey responses were used for analysis, representing 50.6 percent of the total sample population. After removing 4 outliers that took over an hour to complete the survey, average completion time was found to be 6 minutes and 45 seconds. Raw survey data is available in S2 Survey Data.

Qualitative questions were bucketed into numbers ranging from 0-5 (IE 0 to 5 years = 1; 5 to 10 years = 2; etc.). A full set of questions, response bucketing, and UTAUT constructs are included the S1 Appendix A.2. Summary data surrounding survey responses used in the analysis are listed in table 2.

**Table 2.** Summary data for Likert scale questions in survey responses, generated using SmartPLS v. 3.2.9.

| Question | Mean | Median | Min | Max | Standard<br>Deviation | Excess<br>Kurtosis | Skewness |
| --- | --- | --- | --- | --- | --- | --- | --- |
| **Years Practiced – Q1** | 2.326 | 2 | 1 | 5 | 1.434 | -0.932 | 0.659 |
| **Years Old – Q2** | 2.523 | 2 | 1 | 5 | 0.937 | -0.512 | 0.32 |
| **Radiologist Tools – Q4** | 2.326 | 2 | 1 | 5 | 0.982 | -0.162 | 0.426 |
| **Consumer IM – Q5** | 2.488 | 2 | 1 | 5 | 0.774 | 0.556 | 1.189 |
| **Clinical IM – Q6** | 3.023 | 3 | 1 | 5 | 1 | -0.606 | 0.379 |
| **Conversational Agents – Q7** | 3.128 | 3 | 1 | 5 | 1.076 | -0.632 | 0.481 |
| **PE1 – Q11** | 3.733 | 4 | 2 | 5 | 0.813 | -0.354 | -0.259 |
| **EOU1 – Q12** | 3.919 | 4 | 2 | 5 | 0.554 | 1.57 | -0.452 |
| **AF1 – Q13** | 3.733 | 4 | 2 | 5 | 0.722 | -0.22 | -0.106 |
| **ANX1 – Q14** | 2.733 | 3 | 1 | 5 | 1.083 | -0.859 | 0.275 |
| **BI1 – Q15** | 3.872 | 4 | 2 | 5 | 0.679 | 0.219 | -0.289 |
| **PE2 – Q16** | 3.849 | 4 | 2 | 5 | 0.691 | 0.467 | -0.434 |
| **AF2 – Q17** | 3.791 | 4 | 2 | 5 | 0.779 | -0.098 | -0.36 |
| **EOU2 – Q18** | 3.709 | 4 | 2 | 5 | 0.68 | 0.068 | -0.235 |
| **ANX2 – Q19** | 2.465 | 2 | 1 | 5 | 0.936 | -0.377 | 0.665 |
| **PE3 – Q20** | 3.279 | 3 | 1 | 5 | 0.83 | 0.188 | -0.321 |
| **BI2 – Q21** | 3.837 | 4 | 2 | 5 | 0.626 | 1.44 | -0.731 |
| **ANX3 – Q22** | 2.663 | 2 | 1 | 5 | 0.96 | -0.86 | 0.247 |

Partial Least Squares (PLS) Structured Equation Modeling (SEM) was utilized to investigate the relationship between constructs. PLS-SEM calculations were performed using SmartPLS V. 3.2.9. Complete data analysis steps are included in the Supplemental Data Analysis (S1 Appendix A.3). SEM began with connecting all possible paths, then eliminating construct relationships that were insignificant. The final SEM is presented in Fig 3 and details in table 3. T statistics for each path are greater than 1.95 and p values are below 0.05, indicating that each relationship is statistically significant.

**Fig 3.**
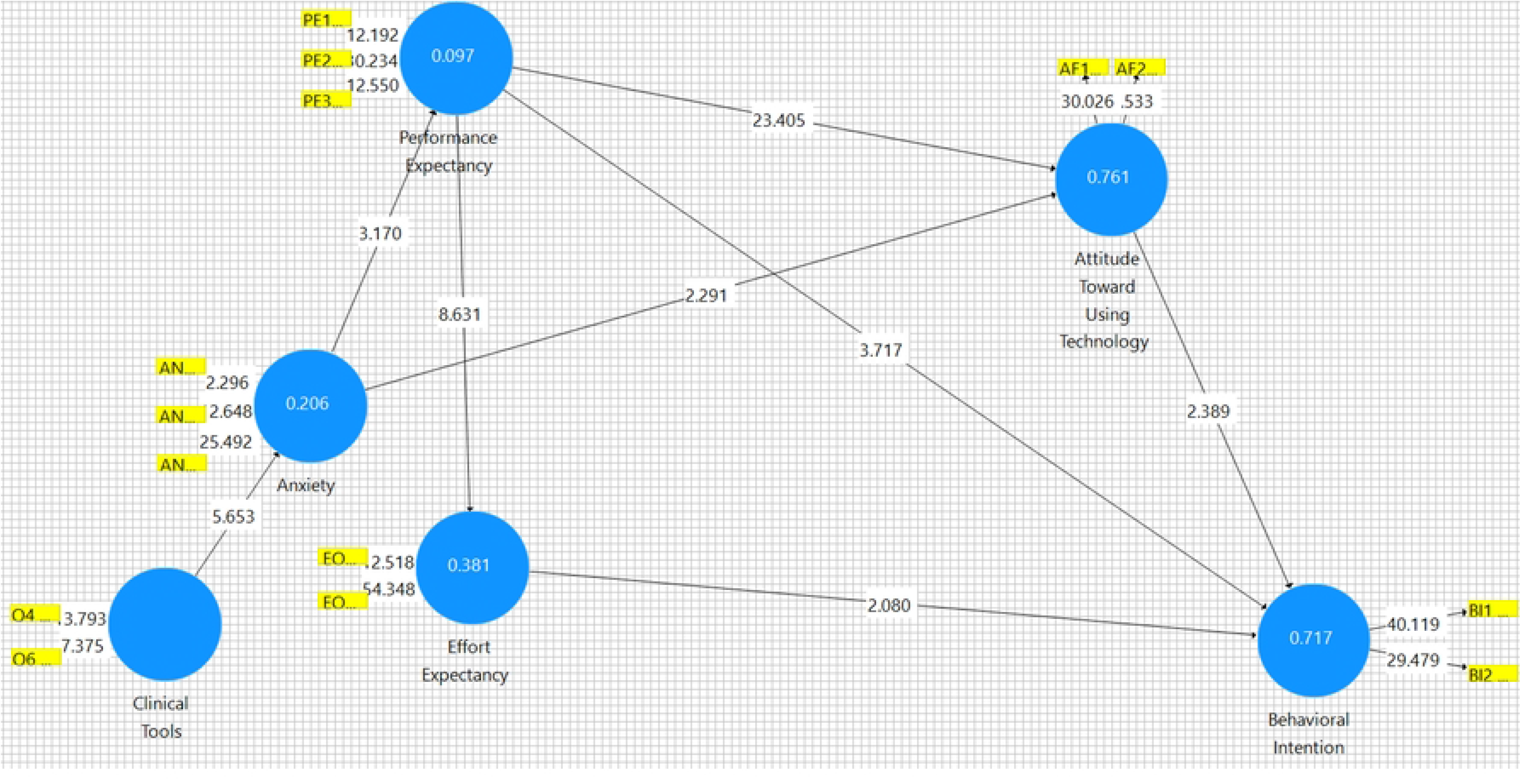
Final Path Model generated using SmartPLS v. 3.2.9 Bootstrapping.

**Table 3.** Final Path Coefficient Report generated using SmartPLS v. 3.2.9 Bootstrapping.

|  | Original<br>Sample<br>(O) | Sample<br>Mean<br>(M) | Standard<br>Deviation<br>(STDEV) | T Statistics<br>(\|O/STDEV\|) | P<br>Values |
| --- | --- | --- | --- | --- | --- |
| **Anxiety -> Attitude Toward Using Technology** | -0.127 | -0.127 | 0.056 | 2.291 | 0.022 |
| **Anxiety -> Performance Expectancy** | -0.312 | -0.326 | 0.098 | 3.17 | 0.002 |
| **Attitude Toward Using Technology -> Behavioral Intention** | 0.329 | 0.326 | 0.138 | 2.389 | 0.017 |
| **Clinical Tools -> Anxiety** | 0.453 | 0.464 | 0.08 | 5.653 | 0 |
| **Effort Expectancy -> Behavioral Intention** | 0.202 | 0.194 | 0.097 | 2.08 | 0.038 |
| **Performance Expectancy -> Attitude Toward Using Technology** | 0.824 | 0.824 | 0.035 | 23.405 | 0 |
| **Performance Expectancy -> Behavioral Intention** | 0.399 | 0.408 | 0.107 | 3.717 | 0 |
| **Performance Expectancy -> Effort Expectancy** | 0.617 | 0.619 | 0.072 | 8.631 | 0 |

Table 4 Cronbach’s Alpha report shows that the t statistic is greater than double the standard deviation, and this indicates the model fits 95% of the data. Table 5 Average Variance Extracted additionally shows strong model fit. Fig 4 Partial Least Squares model was created to determine path coefficients – table 6, and construct validity – table 7.

**Fig 4.**
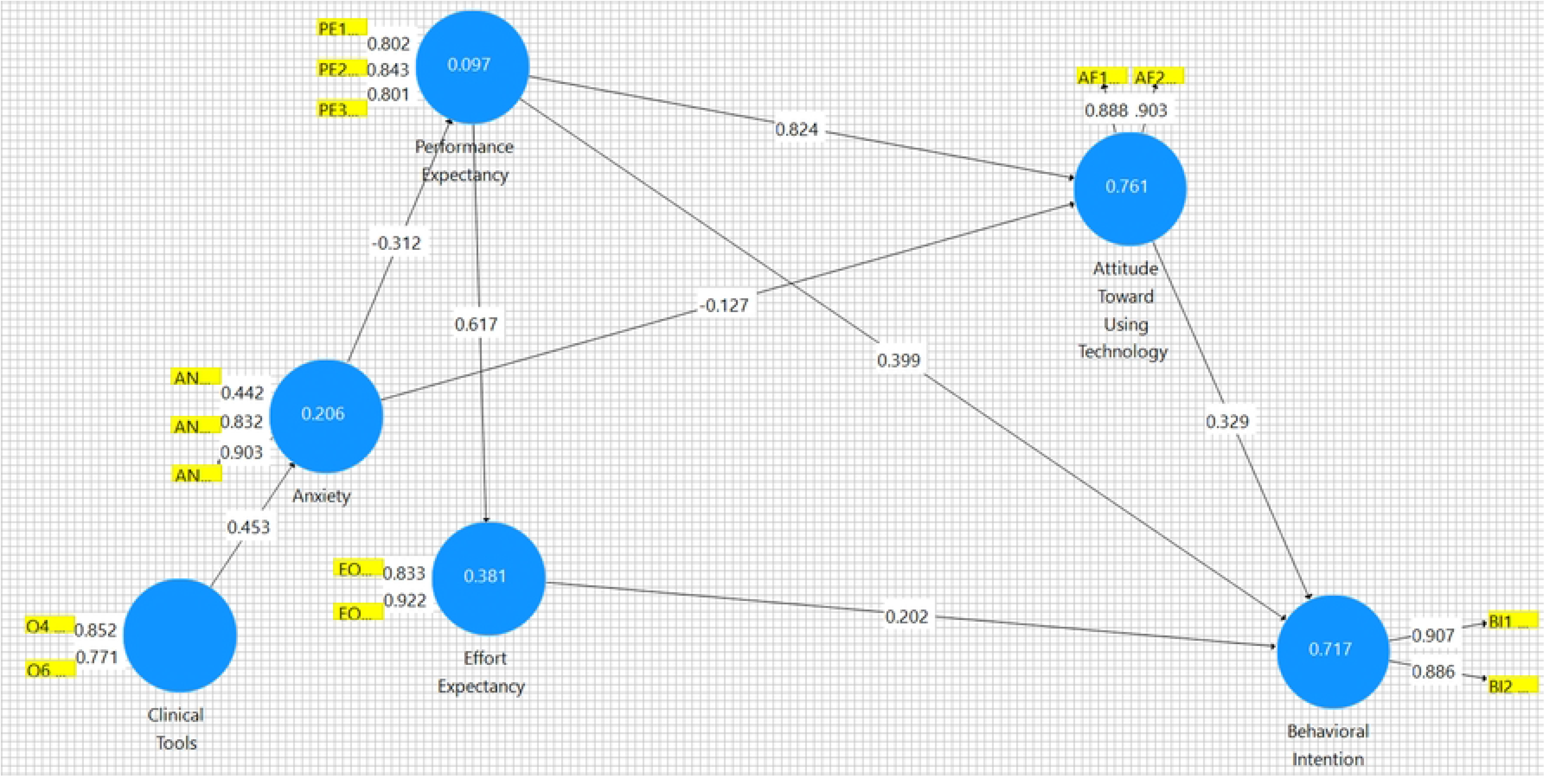
Partial Least Squares Path Model generated using SmartPLS v. 3.2.9 PLS.

**Table 4.**
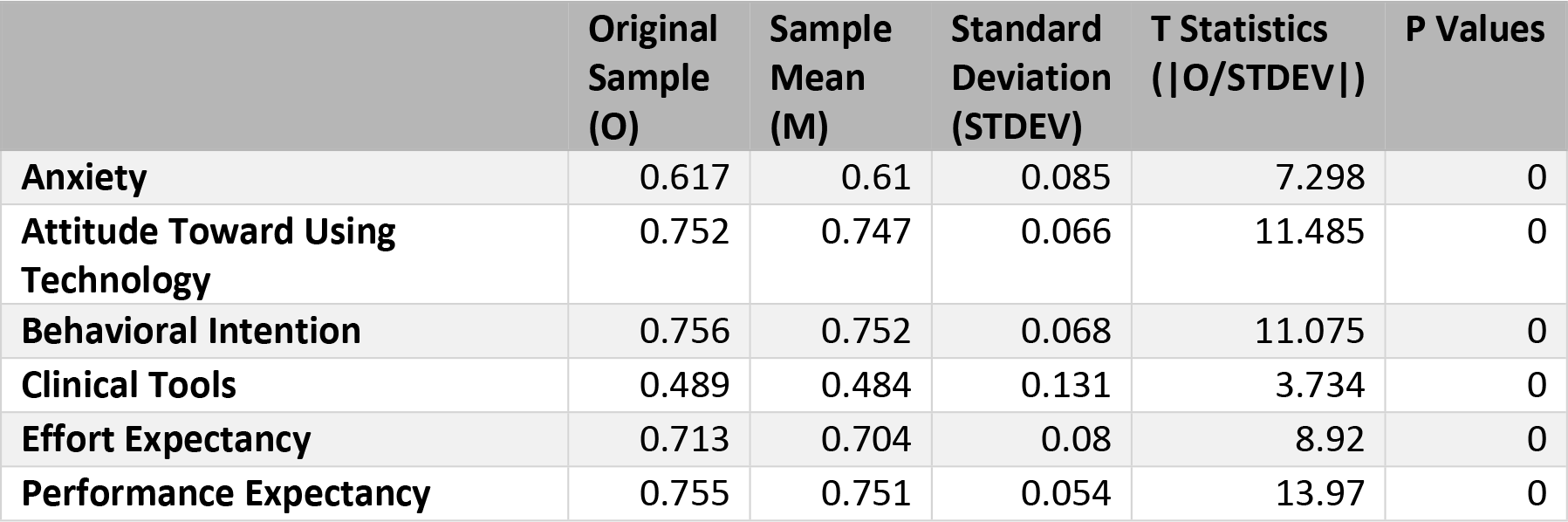
Cronbach’s Alpha Report generated using SmartPLS v. 3.2.9 Bootstrapping.

**Table 5.** Average Variance Extracted (AVE) Report generated using SmartPLS v. 3.2.9 Bootstrapping.

|  | Original<br>Sample<br>(O) | Sample<br>Mean<br>(M) | Standard<br>Deviation<br>(STDEV) | T Statistics<br>(\|O/STDEV\|) | P Values |
| --- | --- | --- | --- | --- | --- |
| Anxiety | 0.568 | 0.569 | 0.051 | 11.249 | 0 |
| Attitude Toward Using<br>Technology | 0.801 | 0.8 | 0.041 | 19.594 | 0 |
| Behavioral Intention | 0.804 | 0.803 | 0.043 | 18.614 | 0 |
| Clinical Tools | 0.66 | 0.659 | 0.058 | 11.467 | 0 |
| Effort Expectancy | 0.772 | 0.767 | 0.051 | 15.171 | 0 |
| Performance Expectancy | 0.665 | 0.665 | 0.05 | 13.394 | 0 |

**Table 6.** Partial Least Squares Path Coefficients Report generated using SmartPLS v. 3.2.9 PLS.

|  | **Anxiety** | **Attitude Toward**<br>**Using Technology** | **Behavioral**<br>**Intention** | **Clinical**<br>**Tools** | **Effort**<br>**Expectancy** | **Performance**<br>**Expectancy** |
| --- | --- | --- | --- | --- | --- | --- |
| **Anxiety** |  | -0.127 |  |  |  | -0.312 |
| **Attitude Toward Using**<br>**Technology** |  |  | 0.329 |  |  |  |
| **Behavioral Intention** |  |  |  |  |  |  |
| **Clinical Tools** | 0.453 |  |  |  |  |  |
| **Effort Expectancy** |  |  | 0.202 |  |  |  |
| **Performance Expectancy** |  | 0.824 | 0.399 |  | 0.617 |  |

**Table 7.** Partial Least Squares Construct Reliability and Validity Report generated using SmartPLS v. 3.2.9 PLS.

|  | **Cronbach's**<br>**Alpha** | **rho_A** | **Composite**<br>**Reliability** | **Average Variance**<br>**Extracted (AVE)** |
| --- | --- | --- | --- | --- |
| **Anxiety** | 0.617 | 0.768 | 0.786 | 0.568 |
| **Attitude Toward Using Technology** | 0.752 | 0.755 | 0.89 | 0.801 |
| **Behavioral Intention** | 0.756 | 0.761 | 0.891 | 0.804 |
| **Clinical Tools** | 0.489 | 0.501 | 0.795 | 0.66 |
| **Effort Expectancy** | 0.713 | 0.777 | 0.871 | 0.772 |
| **Performance Expectancy** | 0.755 | 0.784 | 0.856 | 0.665 |

These final sets of reports explain the model and variance encountered in the model. The weakest relationships surround ANX. Based on this analysis, we know that Clinical Tools strongly influences ANX, however, Clinical Tools has the lowest Cronbach’s Alpha and Adjusted Rho of all reviewed items. ANX also has a less than ideal Cronbach’s Alpha, but other indicators show that it is likely a reliable concept.

### 3.2 Interview Data Analysis

The average interview time was 39.93 minutes. Krippendorff’s CU Alpha was generated at an individual narrative (S1 Appendix B.3) and overall level. Interviews were eliminated until the overall level reached α ≥ .8, resulting in α = 0.82. Code co-occurrence was measured by hypothesis and Sankey diagrams generated (S1 Appendix B.4).

### 3.3 Survey Results

Table 8 includes the outcomes of each hypothesis for the survey. Results are expanded upon in S1 Appendix A.4.

**Table 8.** Survey Hypotheses tested, P-values, and outcomes.

| Hypothesis | P-Value | Reject Null |
| --- | --- | --- |
| **H1: Radiologists have intent to use an IM based conversational agent** | <0.001 | Yes |
| H1-A: Intent is moderated by performance expectancies | <0.001 | Yes |
| H1-B: Intent is moderated by effort expectancies | <0.001 | Yes |
| H1-C: Intent is moderated by anxiety | 0.514 | No |
| H1-D: Intent is moderated by age | 0.103 | No |
| H1-E: Intent is moderated by radiologist's experience with general consumer conversational agents and experience with radiology domain specific clinical tools | 0.336 | No |
| H1-F: Intent is moderated by attitude toward the system | 0.017 | Yes |
| **H2: Radiologists attitude toward the IM based conversational agent is positive** | <0.001 | Yes |
| H2-A: Attitude is moderated by performance expectancies | <0.001 | Yes |
| H2-B: Attitude is moderated by effort expectancies | 0.056 | No |
| H2-C: Attitude is moderated by anxiety | 0.022 | Yes |
| H2-D: Attitude is moderated by age | 0.103 | No |
| H2-E: Attitude is moderated by radiologist's experience with general consumer conversational agents and experience with radiology domain specific clinical tools | 0.371 | No |
| **H3: Age influences radiologist's perspective of the intervention** | Multiple | No |
| **H4: Experience with consumer conversational agent's moderate's radiologist's perspective of the intervention** | Multiple | No |
| **H5: Experience with consumer and clinical IM tools moderate's radiologist's perspective of the intervention.** | Multiple | No |
| **H6: Experience with radiology domain specific clinical tools moderate's radiologist's perspective of the intervention.** | Multiple | Yes |

Hypotheses were tested at a 95% significance level.

### 3.4 Interview Results

Table 9 includes the outcomes of each hypothesis for the interview. Interpretation, code co-occurrence tables, and Sankey diagrams supporting results are found in S1 Appendix B.4.

**Table 9.** Interview Hypotheses tested.

| Hypothesis | Reject Null |
| --- | --- |
| H1: Expected effort is not a contributing factor in attitude towards the intervention but is a contributing factor in intent to use the intervention. | No |
| **H2: With respect to this intervention, anxiety has a negative relationship with performance expectancy. As anxiety increases, performance expectancy decreases. As anxiety decreases, performance expectancy increases.** | No |
| **H3: Radiologists have a positive attitude towards this intervention and a high intent to use this intervention if it were produced.** | Yes |
| **H4: For this intervention, intent to use and attitude are mostly influenced by performance expectancies.** | No |
| H4-A: Radiologist's attitude towards this intervention is mostly influenced by the expected performance of the system. | No |
| H4-B: Radiologist's intent to use this intervention is mostly influenced by expected performance of the system. | No |
| **H5: Radiologist's performance expectancies positively influence their effort expectancies. As expected performance increases, expected effort decreases.** | Yes |

## 5 Discussion

Radiologists have a high intent to use and positive attitudes towards IM based CDSS and the presented interventions overall. We determined that years of experience, and Consumer Tools (IM and CA) were not moderating variables in our model. In any given path, the t statistic was too low and p value too high to consider this in our analysis. These questions are not a part of the UTAUT model, and we found them not to be factors relevant to our efforts. The following UTAUT expected paths were additionally removed, and speculation as to why is included:

### 5.1 Age and Intent to Use

This is a deviation from UTAUT. Potentially, radiologists are technologically saturated users; they perform their work functions using a wide variety of complex technological solutions. Among clinicians, radiologists chose this specialty because of their interest in technology solutions within healthcare. We were unable to measure this result during interviews.

### 5.2 Expected Efforts Influence on Attitude

The survey and interview studies have opposing results for expected efforts influence on attitude. The survey deviated from UTAUT in not finding an association with attitude. However, the interview study showed that decreasing effort is linked to positive attitude and positive intent to use. Common themes on effort/attitude interactions-

- Reducing time to acquire and apply clinical knowledge.
  - “…however many seconds it takes for everyone…and it’s different for everyone to figure out how they want to go about finding this information. We all kind of I think most people know where to look IE the ACR guidelines, but…having this thought process kind of forcing us to focus on this dialog box kind of streamlines that whole process. So I think overall, it should enhance the workflow”
  - “…these are the things when we’re not given enough information…Some people perseverate on the lack of information more than others. And some people are really dutiful and want to go into the EMR and look, but that could be one to two minutes, and then compound that over an entire shift. Do that a couple times. That could be an hour that you’ve saved if you had this information in a ready format, or in a readily available format, so I think this definitely makes you, from this particular type of interactions definitely makes you more efficient, I believe.”
- Increasing multitasking
  - “I think it is a great idea. I think it helps you do multiple things, maybe not just in this cardiac workup. But like for lung nodules when your kind of trying to decide what the appropriate workup is. We always have a caveat that takes seconds to say but it’s still seconds that you have to say it every time. You know, if the patient has high risk for pulmonary malignancy recommend whatever. We know that they’re already high risk for whatever, then I feel like we don’t need to say that. Or if that even auto populates the patient has these risk factors that we would recommend discussing these risk factors.”
- Trusting CDSS as safety nets
  - “…we touted on AI is not to replace your diagnostic skills, but eight other things, whether it’s making you more efficient or providing kind of a little safety net, right. Maybe you forgot to mention a follow up or something that really should be a critical result.”

### 5.3 Anxieties influence on Attitude

The survey shows a small negative relationship with attitude. The interview study asked many questions to understand anxiety surrounding this intervention, however, we were unable to strongly correlate with attitude. Overall, anxiety is the least grounded concept throughout the interview.

### 5.4 Expected Performance as the Major Influencer of Attitude and Intent to Use

Overall, expected performance is a major influence on attitude and intent to use. Within the survey results it has significantly more influence than any other factor. However, the interview results show a stronger correlation of expected effort with attitude and intent to use. There is a strong negative relationship between performance and effort present in both phases of the study, another deviation from the UTAUT model. There is potential that radiologists’ system use is derived from performance, maybe measured in clinical outcomes. However, we cannot assume these performance metrics overcome effort needs. Common themes from factors influencing attitude/intent to use-

- Radiologists expect to be interrupted or context switch quickly.
  - “Interrupt My normal workflow? Well, I guess it depends on what is normal. This would not interrupt my normal workflow. We’re constantly getting interrupted. It would just be another interruption among a series of normal interruptions.”
  - “…this is kind of the thought process that I, this is I go through this checklist. Basically, every time I close a study, we look at the work list again. I’m thinking to myself, looking over my shoulder at the residents and looking at their, their work list, and thinking about [county hospital] over here, looking at how deep my work list is how far so I basically run this checklist mentally, in between each exam.”
- Reducing effort is highly embraced.
  - “I love the idea that I’m not having to call someone and that automatically reminds me and I can just either do one click and go one click would be nice to just be like, Yes.”
  - “the status quo is quicker or definitely is quicker than then that interface I just saw on the video.”
  - “…I’m responsible for all those things that I don’t see on a regular basis…Let me go back to that algorithm figure out what I need to say. This would be a really great tool for me in those cases, because I don’t have to worry as much I think I’m missing a recommendation or something like that. I don’t have to hedge as much. I don’t have to hurry try to get to get to my list. Luckily, we don’t aren’t too inundated so it’s not an issue but I do feel like this will help me to put the appropriate things in with the appropriate recommended”
  - “Well, it would shortcut having to call a technologist and initiate a conversation about what the patient was, what the study was, the post processing that you needed, done. So if the radbot could predict that you might need it. And could figure out what you needed quicker and would negate having that phone call that would be positive.”
- Radiologists will trade effort for performance.
  - “So it slows you down slightly, but in the long run of collections and all that stuff. Yeah, I think it would [improve performance], because you’re making sure you get reimbursed and get the correct RVU amounts for the right study.”
  - “… the amount of time it takes to look those things up, and it’s not super frequent, but not super infrequent…either it’s taking you more time to figure it out or you also end up with more a variation amongst different radiologists for the recommendation. So you’re not only might save time, but you might also decrease the heterogeneity of the recommendations. And probably more, you’d be probably more likely to actually be following the guidelines since you’d be prompted to to adhere to them.”
  - “…I think maybe it slows you slightly on the front end, but on the back end, it helps you, and it helps clinicians too”
  - “I think it would decrease my productivity very minimally. But for a good cause.”

### 5.5 Conclusion

“No, I think again, this is it’s all these videos have demonstrated processes which are mostly done mentally by all radiologists. And again, it’s not always easy to kind of put these things on a screen or you know, because your kind of you’re, you’re juggling a couple different priorities at the same time. So, I think this is kind of taking an existing thing and it’s making a more organized and streamlined fashion.”

Radiologist’s interactions with decision support tools, or at least this intervention, differs from the standard user software interaction model. The positive relationship from performance to effort is the most major deviation, allowing increasing effort if the outcomes are desirable enough. This relationship is supported by both the survey and interview studies. Further, because performance and effort make up most of attitude and intent to use, there are a lot of opportunities for CDSS to provide novel workflow changes that increase patient outcomes. CDSS should be designed to streamline activities, and we see particular interest in tools to enable clinical knowledge gathering and context switching.

“Yeah, assuming that you would have gone to the EMR it was important enough to go there. And if you went there to take more time, okay. But at the same point, you know, it might change the threshold at which you would ask a question, right? It’s like, it’d be nice if we knew this and it’s easy just to query it. But you otherwise might not go to the EMR.”

Anxiety is another large deviation from the standard user model. In both parts of the study anxiety had the weakest relationships and was often secondary to the excitement of new clinical solutions. The most common source of anxiety surrounds the maintenance of CDSS “I guess the part that causes me to pause is who’s going to be mining for new updates? And how can we be sure that we’re staying current on recommendations? You know, how is that? Who’s going to handle that part of it?”

Radiologists deviate from the standard clinician with regards to the 10 commandments of CDSS. Commandments 2, 3, 7, 10, and 1 – anticipate needs, fit into user workflow, simple interventions, knowledge system maintenance, and speed – are all highlighted within radiology specific guidance, and we do find these present for radiologists in our study. However, the relationship between performance and effort highlights that radiologist CDSS doesn’t need to always hit every commandment. Radiologists expect workflow modification, they routinely use complex interventions, and they are not overwhelmed by CDSS information gathering. As we design for the future radiologist, we can trade effort in these commandments for increasing positive outcomes.

## Data Availability

All relevant survey data are within the manuscript and its Supporting Information files. Interview data is withheld due to IRB restrictions.

## Supporting Information Captions

**S1 Appendix.** Detailed information on the study including the full survey, expanded hypothesis results, Semi-structured interview guide, code co-occurrence tables, full data analysis/findings, and additional diagrams.

**S2 Survey Data.** Raw quantitative survey data in Excel format.

## Notes

### Competing Interest Statement

The authors have declared no competing interest.

### Funding Statement

John Burns - No disclosures or competing interests. Marc Kohli - Travel support from SIIM and RSNA during the study period. Co-Founder and Shareholder in Alara Imaging, which was not involved in the study and sells products related to quality measures and edge-to-cloud gateways. Josette Jones - No disclosures or competing interests. Saptarshi Purkayastha and Judy W. Gichoya are funded by US National Science Foundation (grant number 1928481) from the Division of Electrical, Communication & Cyber Systems. Judy W. Gichoya is also funded by the National Institute of Biomedical Imaging and Bioengineering (NIBIB) MIDRC grant of the National Institutes of Health (75N92020C00008 and 75N92020C00021). Saptarshi Purkayastha has no competing interests with regards to this work. Judy W. Gichoya has no competing interests with regards to this work.

### Author Declarations

Indiana University IRB https://research.iu.edu/forms/human-subjects-irb.html

## References

1 Choy G, Khalilzadeh O, Michalski M, Do S, Samir AE, Pianykh OS, et al. Current Applications and Future Impact of Machine Learning in Radiology. Radiology. 2018 Aug;288(2):318–28.

2 Dreyer KJ, Geis JR. When Machines Think: Radiology’s Next Frontier. Radiology. 2017 Dec;285(3):713–8.

3 Gichoya JW, Alarifi M, Bhaduri R, Tahir B, Purkayastha S. Using cognitive fit theory to evaluate patient understanding of medical images. Annual International Conference of the IEEE Engineering in Medicine and Biology Society IEEE Engineering in Medicine and Biology Society Annual International Conference [Internet]. 2017 Jul 1 [cited 2023 Mar 23];2017:2430–3. Available from: https://pubmed.ncbi.nlm.nih.gov/29060389/

4 Gichoya J, Nuthakki S, Maity PG, Purkayastha S. Phronesis of AI in radiology: Superhuman meets natural stupidity. ArXiv [Internet]. 2018 [cited 2023 Mar 23]; Available from: https://www.semanticscholar.org/paper/Phronesis-of-AI-in-radiology%3A-Superhuman-meets-Gichoya-Nuthakki/57e870688e760fe04f5b2db3ddd5d42897cc2d0d

5 Venkatesh V, Morris MG, Davis GB, Davis FD. User Acceptance of Information Technology: toward a Unified View. MIS Quarterly [Internet]. 2003;27(3):425–78. Available from: https://www.jstor.org/stable/pdf/30036540

6 Lodwick GS, Turner AH, Lusted LB, Templeton AW. Computer-aided analysis of radiographic images. Journal of Chronic Diseases [Internet]. 1966 Apr 1 [cited 2023 Mar 23];19(4):485–96. Available from: https://www.sciencedirect.com/science/article/abs/pii/0021968166901226

7 Agrawal JP, Erickson BJ, Kahn CE. Imaging Informatics: 25 Years of Progress. Yearbook of Medical Informatics [Internet]. 2016 May 20;(Suppl 1):S23–31. Available from: https://www.ncbi.nlm.nih.gov/pmc/articles/PMC5171495/

8 Nowinski WL, Qian G, Hanley DF. A CAD System for Hemorrhagic Stroke. The Neuroradiology Journal. 2014 Aug;27(4):409–16.

9 Stivaros SM, Gledson A, Nenadic G, Zeng X-J, Keane J, Jackson A. Decision support systems for clinical radiological practice — towards the next generation. The British Journal of Radiology. 2010 Nov;83(995):904–14.

10 Wang Y, Yan F, Lu X, Zheng G, Zhang X, Wang C, et al. IILS: Intelligent imaging layout system for automatic imaging report standardization and intra-interdisciplinary clinical workflow optimization. EBioMedicine. 2019 Jun;44:162–81.

11 L. Barinov et al., "Impact of Data Presentation on Physician Performance Utilizing Artificial Intelligence-Based Computer-Aided Diagnosis and Decision Support Systems," (in eng), J Digit Imaging, vol. 32, no. 3, pp. 408-416, Jun 2019, doi: 10.1007/s10278-018-0132-5.

12 K. S. Berbaum and E. A. Franken, Jr., "Commentary does clinical history affect perception?," (in eng), Acad Radiol, vol. 13, no. 3, pp. 402-3, Mar 2006, doi: 10.1016/j.acra.2005.11.031.

13 K. S. Berbaum, E. A. Franken, Jr., D. D. Dorfman, and K. R. Lueben, "Influence of clinical history on perception of abnormalities in pediatric radiographs," (in eng), Acad Radiol, vol. 1, no. 3, pp. 217-23, Nov 1994, doi: 10.1016/s1076-6332(05)80717-2.

14 A. Leslie, A. J. Jones, and P. R. Goddard, "The influence of clinical information on the reporting of CT by radiologists," Br J Radiol, vol. 73, no. 874, pp. 1052-1055, 2000, doi: 10.1259/bjr.73.874.11271897.

15 B. I. Reiner, "Medical Imaging Data Reconciliation, Part 3: Reconciliation of Historical and Current Radiology Report Data," Journal of the American College of Radiology, vol. 8, no. 11, pp. 768-771, 2011/11/01/ 2011, doi: https://doi.org/10.1016/j.jacr.2011.04.021.

16 R. J. Gorniak et al., "A PACS-Integrated Tool to Automatically Extract Patient History From Prior Radiology Reports," (in eng), J Am Coll Radiol, vol. 13, no. 10, pp. 1249-1252, Oct 2016, doi: 10.1016/j.jacr.2016.06.004.

17 G. W. Boland et al., "Decision support for radiologist report recommendations," (in eng), J Am Coll Radiol, vol. 8, no. 12, pp. 819-23, Dec 2011, doi: 10.1016/j.jacr.2011.08.003.

18 B. H. Do, A. S. Wu, J. Maley, and S. Biswal, "Automatic retrieval of bone fracture knowledge using natural language processing," (in eng), J Digit Imaging, vol. 26, no. 4, pp. 709-13, Aug 2013, doi: 10.1007/s10278-012-9531-1.

19 M. Kohli et al., "Bending the Artificial Intelligence Curve for Radiology: Informatics Tools From ACR and RSNA," (in eng), J Am Coll Radiol, Jul 15 2019, doi: 10.1016/j.jacr.2019.06.009.

20 Y. Liu, L. N. Zhu, Q. Liu, C. Han, X. D. Zhang, and X. Y. Wang, "Automatic extraction of imaging observation and assessment categories from breast magnetic resonance imaging reports with natural language processing," (in eng), Chin Med J (Engl), vol. 132, no. 14, pp. 1673–1680, Jul 20 2019, doi: 10.1097/cm9.0000000000000301.

21 M. Esmaeili, S. M. Ayyoubzadeh, N. Ahmadinejad, M. Ghazisaeedi, A. Nahvijou, and K. Maghooli, "A decision support system for mammography reports interpretation," (in eng), Health Inf Sci Syst, vol. 8, no. 1, p. 17, Dec 2020, doi: 10.1007/s13755-020-00109-5.

22 S. Bozkurt, F. Gimenez, E. S. Burnside, K. H. Gulkesen, and D. L. Rubin, "Using automatically extracted information from mammography reports for decision-support," (in eng), Journal of biomedical informatics, vol. 62, pp. 224-31, Aug 2016, doi: 10.1016/j.jbi.2016.07.001.

23 T. A. Patel et al., "Correlating mammographic and pathologic findings in clinical decision support using natural language processing and data mining methods," (in eng), Cancer, vol. 123, no. 1, pp. 114–121, Jan 1 2017, doi: 10.1002/cncr.30245.

24 R. European Society of, "The future role of radiology in healthcare," (in eng), Insights Imaging, vol. 1, no. 1, pp. 2-11, 2010, doi: 10.1007/s13244-009-0007-x.

25 D. L. Weiss, W. Kim, B. F. t. Branstetter, and L. M. Prevedello, "Radiology reporting: a closed-loop cycle from order entry to results communication," (in eng), J Am Coll Radiol, vol. 11, no. 12 Pt B, pp. 1226-37, Dec 2014, doi: 10.1016/j.jacr.2014.09.009.

26 P. A. Larson, L. L. Berland, B. Griffith, C. E. Kahn, Jr., and L. A. Liebscher, "Actionable findings and the role of IT support: report of the ACR Actionable Reporting Work Group," (in eng), J Am Coll Radiol, vol. 11, no. 6, pp. 552-8, Jun 2014, doi: 10.1016/j.jacr.2013.12.016.

27 X. Meng, C. H. Ganoe, R. T. Sieberg, Y. Y. Cheung, and S. Hassanpour, "Assisting radiologists with reporting urgent findings to referring physicians: A machine learning approach to identify cases for prompt communication," (in eng), Journal of biomedical informatics, vol. 93, p. 103169, May 2019, doi: 10.1016/j.jbi.2019.103169.

28 R. Lacson et al., "Four-year impact of an alert notification system on closed-loop communication of critical test results," (in eng), AJR Am J Roentgenol, vol. 203, no. 5, pp. 933-8, Nov 2014, doi: 10.2214/ajr.14.13064.

29 A. B. Rosenkrantz, J. Sherwin, C. P. Prithiani, D. Ostrow, and M. P. Recht, "Technology-Assisted Virtual Consultation for Medical Imaging," (in eng), J Am Coll Radiol, vol. 13, no. 8, pp. 995-1002, Aug 2016, doi: 10.1016/j.jacr.2016.02.029.

30 B. I. Reiner, "Redefining the Practice of Peer Review Through Intelligent Automation-Part 3: Automated Report Analysis and Data Reconciliation," (in eng), J Digit Imaging, vol. 31, no. 1, pp. 1-4, Feb 2018, doi: 10.1007/s10278-017-0006-2.

31 B. I. Reiner, "Quantifying Analysis of Uncertainty in Medical Reporting: Creation of User and Context-Specific Uncertainty Profiles," (in eng), J Digit Imaging, vol. 31, no. 4, pp. 379-382, Aug 2018, doi: 10.1007/s10278-018-0057-z.

32 J. L. Burns, D. Hasting, J. W. Gichoya, B. McKibben, 3rd, L. Shea, and M. Frank, "Just in Time Radiology Decision Support Using Real-time Data Feeds," (in eng), J Digit Imaging, Sep 12 2019, doi: 10.1007/s10278-019-00268-2.

33 R. Chen, P. Mongkolwat, and D. S. Channin, "RadMonitor: radiology operations data mining in real time," (in eng), J Digit Imaging, vol. 21, no. 3, pp. 257-68, Sep 2008, doi: 10.1007/s10278-007-9033-8.

34 J. W. Nance, Jr., C. Meenan, and P. G. Nagy, "The future of the radiology information system," (in eng), AJR Am J Roentgenol, vol. 200, no. 5, pp. 1064-70, May 2013, doi: 10.2214/ajr.12.10326.

35 P. G. Nagy, M. J. Warnock, M. Daly, C. Toland, C. D. Meenan, and R. S. Mezrich, "Informatics in radiology: automated Web-based graphical dashboard for radiology operational business intelligence," (in eng), Radiographics: a review publication of the Radiological Society of North America, Inc, vol. 29, no. 7, pp. 1897-906, Nov 2009, doi: 10.1148/rg.297095701.

36 M. B. Morgan, B. F. t. Branstetter, D. M. Lionetti, J. S. Richardson, and P. J. Chang, "The radiology digital dashboard: effects on report turnaround time," (in eng), J Digit Imaging, vol. 21, no. 1, pp. 50-8, Mar 2008, doi: 10.1007/s10278-007-9008-9.

37 O. A. Awan, F. van Wagenberg, M. Daly, N. Safdar, and P. Nagy, "Tracking delays in report availability caused by incorrect exam status with Web-based issue tracking: a quality initiative," (in eng), J Digit Imaging, vol. 24, no. 2, pp. 300-7, Apr 2011, doi: 10.1007/s10278-010-9330-5.

38 H. International. "HL7 International." http://www.hl7.org/ (accessed 9/23/2019, 2019).

39 D. Library. "About DICOM." https://www.dicomlibrary.com/dicom/ (accessed 9/23/2019, 2019).

40 M. Kohli, K. J. Dreyer, and J. R. Geis, "Rethinking Radiology Informatics," American Journal of Roentgenology, vol. 204, no. 4, pp. 716–720, 2015/04/01 2015, doi: 10.2214/AJR.14.13840.

41 D. Teather, B. A. Morton, G. H. du Boulay, K. M. Wills, D. Plummer, and P. R. Innocent, "Computer assistance for C.T. scan interpretation and cerebral disease diagnosis," (in eng), Stat Med, vol. 4, no. 3, pp. 311–5, Jul-Sep 1985, doi: 10.1002/sim.4780040310.

42 R. Khorasani, "Clinical decision support in radiology: what is it, why do we need it, and what key features make it effective?," (in eng), J Am Coll Radiol, vol. 3, no. 2, pp. 142-3, Feb 2006, doi: 10.1016/j.jacr.2005.11.008.

43 D. W. Bates et al., "Ten commandments for effective clinical decision support: making the practice of evidence-based medicine a reality," (in eng), Journal of the American Medical Informatics Association: JAMIA, vol. 10, no. 6, pp. 523–30, Nov-Dec 2003, doi: 10.1197/jamia.M1370.

44 S. E. Cheeseman, "Communication and collaboration technologies," (in eng), Neonatal Netw, vol. 31, no. 2, pp. 115–9, Mar-Apr 2012, doi: 10.1891/0730-0832.31.2.115.

45 C. Pimmer, S. Mhango, A. Mzumara, and F. Mbvundula, "Mobile instant messaging for rural community health workers: a case from Malawi," (in eng), Glob Health Action, vol. 10, no. 1, p. 1368236, 2017, doi: 10.1080/16549716.2017.1368236.

46 J. R. Bautista and T. T. C. Lin, "Nurses’ use of mobile instant messaging applications: A uses and gratifications perspective," (in eng), Int J Nurs Pract, vol. 23, no. 5, Oct 2017, doi: 10.1111/ijn.12577.

47 T. B. Iversen, L. Melby, and P. Toussaint, "Instant messaging at the hospital: supporting articulation work?," (in eng), Int J Med Inform, vol. 82, no. 9, pp. 753-61, Sep 2013, doi: 10.1016/j.ijmedinf.2013.05.004.

48 C. Rosset, A. Rosset, and O. Ratib, "General consumer communication tools for improved image management and communication in medicine," (in eng), J Digit Imaging, vol. 18, no. 4, pp. 270-9, Dec 2005, doi: 10.1007/s10278-005-6703-2.

49 L. Fratt, "PACS Powers the Enterprise," Health Imaging Insights in Imaging & Informatics, 10/15/2007. [Online]. Available: https://www.healthimaging.com/topics/advanced-visualization/pacs-powers-enterprise

50 "Philips Adds Options to PACS," Imaging Technology News, 6/11/2007. [Online]. Available: https://www.itnonline.com/content/philips-adds-options-pacs

51 A. Grabb, "Early experience with electronic messaging tightly integrated within PACS," (in eng), J Am Coll Radiol, vol. 8, no. 2, pp. 141-2, Feb 2011, doi: 10.1016/j.jacr.2010.10.007.

52 I. N. America. "INFINITT PACS." https://www.infinittna.com/solutions/radiology/infinitt-pacs/ (accessed 10/21/2019, 2019).

53 I. W. Health. "Merge PACS™ Innovative Reading Workflows for Enterprise Radiology" https://www.merge.com/Solutions/Radiology/Merge-PACS.aspx (accessed 10/21/2019, 2019).

54 I. Carestream Health. RIS Module Streamlined Productivity. (2018). [Online]. Available: https://collaboration.carestream.com/sites/default/files/brochure_ris_module_201810.pdf

55 Saince. "Saince Merge Enterprise PACS." https://www.saince.com/international-solutions/saince-enterprise-pacs/ (accessed 10/21/2019, 2019).

56 S. Medical. "Sectra PACS and RIS - Examples of supported radiology workflows: Communication." https://medical.sectra.com/product/sectra-radiology-pacs-ris/ (accessed 10/21/2019, 2019).

57 F. H. A. Corporation. "Synapse® EIS Features." https://www.fujifilmusa.com/products/medical/medical-informatics/radiology/RIS/index.html#features (accessed 10/21/2019, 2019).

58 A. HealthCare. "XERO Viewer All images, One View." https://global.agfahealthcare.com/us/enterprise-imaging/universal-viewer/ (accessed 10/21/2019, 2019).

59 D. McFarlane, "Comparison of four primary methods for coordinating the interruption of people in human-computer interaction," Hum.-Comput. Interact., vol. 17, no. 1, pp. 63-139, 2002, doi: 10.1207/s15327051hci1701_2.

60 M. Bates, "Health Care Chatbots Are Here to Help," (in eng), IEEE Pulse, vol. 10, no. 3, pp. 12–14, May-Jun 2019, doi: 10.1109/mpuls.2019.2911816.

61 L. Laranjo et al., "Conversational agents in healthcare: a systematic review," (in eng), Journal of the American Medical Informatics Association: JAMIA, vol. 25, no. 9, pp. 1248–1258, Sep 1 2018, doi: 10.1093/jamia/ocy072.

62 M. Beveridge and J. Fox, "Automatic generation of spoken dialogue from medical plans and ontologies," (in eng), Journal of biomedical informatics, vol. 39, no. 5, pp. 482-99, Oct 2006, doi: 10.1016/j.jbi.2005.12.008.

63 B. Mesko, G. Hetenyi, and Z. Gyorffy, "Will artificial intelligence solve the human resource crisis in healthcare?," (in eng), BMC Health Serv Res, vol. 18, no. 1, p. 545, Jul 13 2018, doi: 10.1186/s12913-018-3359-4.

64 S. i. Breastfeeding. "A virtual assistant to help doctors in their daily work." https://www.safeinbreastfeeding.com/safedrugbot-chatbot-medical-assistant/ (accessed 10/27/2019, 2019).

65 A. Gupta, H. Li, and R. Sharda, "Should I send this message? Understanding the impact of interruptions, social hierarchy and perceived task complexity on user performance and perceived workload," Decis. Support Syst., vol. 55, no. 1, pp. 135-145, 2013, doi: 10.1016/j.dss.2012.12.035.

66 M. Czerwinski, E. Cutrell, and E. Horvitz, "Instant Messaging: Effects of Relevance and Timing," 11/20 2000.

67 Qualtrics. "QualtricsXM." https://www.qualtrics.com/ (accessed 11/2/2019, 2019).

68 E. Wiki. "Usability and user experience surveys." http://edutechwiki.unige.ch/en/Usability_and_user_experience_surveys#UTAUT (accessed 11/2/2019, 2019).

69 Sudraben, L. X-ray.jpg, Ed., ed. wikimedia: Wikimedia, 2018.

70 O. A. Imaging, "Jane_Doe_CBCT_NEW_Report," Jane_Doe_CBCT_NEW_Report.jpg, Ed., ed. http://www.orbitimaging.com/imaging-services/radiologist-interpretation/.

71 "screen-0," screen-0.jpg, Ed., ed.

72 W. Hsu, "Capturing Data Elements and the Role of Imaging Informatics." [Online]. Available: http://amos3.aapm.org/abstracts/pdf/99-27434-359478-111844-1383861762.pdf

73 I. Zoom Video Communications. "Zoom." https://zoom.us/ (accessed 8/22/2021, 2021).

74 Otter.AI. "Otter.AI." https://otter.ai (accessed 3/16/2021, 2021).

75 K. Krippendorff, Content Analysis: An Introduction to Its Methodology. Sage, 2004.

76 R. Likert, "A technique for the measurement of attitudes," Archives of Psychology, vol. 22 140, pp. 55–55, 1932.

77 J. Willatt, S. Chong, J. A. Ruma, and J. Kuriakose, "Incidental Adrenal Nodules and Masses: The Imaging Approach," (in eng), Int J Endocrinol, vol. 2015, pp. 410185-410185, 2015, doi: 10.1155/2015/410185.

